# Potential role of fructose on human colon DNA methylation in racial disparities observed for colorectal cancer risk

**DOI:** 10.1101/2023.05.31.23290777

**Authors:** Matthew A. Devall, Stephen Eaton, Gaizun Hu, Xiangqing Sun, Ethan Jakum, Samyukta Venkatesh, Steven M. Powell, Cynthia Yoshida, Daniel J. Weisenberger, Gregory S. Cooper, Joseph Willis, Seham Ebrahim, Jamie Zoellner, Graham Casey, Li Li

## Abstract

**Background and aims:** An increasing body of observational studies has linked fructose intake to colorectal cancer (CRC). African Americans (AAs) are significantly more likely than European Americans to consume greater quantities of fructose and to develop right-side colon cancer. Yet, a mechanistic link between these two associations remains poorly defined. We aimed to identify differentially methylated regions (DMRs) associated with dietary fructose consumption measures obtained from food frequency questionnaires in a cohort of normal colon biopsies derived from AA men and women (n=79)

**Methods:** DNA methylation data from this study was obtained using the Illumina Infinium MethylationEPIC kit and is housed under accession GSE151732. DMR analysis was carried out using *DMRcate* in right and matched left colon, separately. Secondary analysis of CRC tumors was carried out using data derived from TCGA-COAD, GSE101764 and GSE193535. Differential expression analysis was carried out on CRC tumors from TCGA-COAD using *DESeq2*.

**Results:** We identified 4,263 right-side fructose-DMRs. In contrast, only 24 DMRs survived multiple testing corrections (FDR<0.05) in matched, left colon. To identify targets by which dietary fructose drives CRC risk, we overlaid these findings with data from three CRC tumor datasets. Remarkably, almost 50% of right-side fructose-DMRs overlapped regions associated with CRC in at least one of three datasets. *TNXB* and *CDX2* ranked among the most significant fructose risk DMRs in right and left colon respectively that also displayed altered gene expression in CRC tumors.

**Conclusions:** Our mechanistic data support the notion that fructose has a greater CRC-related effect in right than left AA colon, alluding to a potential role for fructose in contributing to racial disparities in CRC.

## Introduction

The natural sweetness of dietary fructose makes it an appealing additive to processed foods and beverages, and most caloric sweeteners contain at least 40% fructose^1^. High fructose corn syrup (HFCS) is the most common form of sweetener added to sugar sweetened beverages (SSB)^2, 3^. Shifts in dietary patterns have seen dramatic increase of HFCS consumption in recent years, with research showing it to account for approximately 6-10% of caloric intake among US adults^4-6^. Similarly, there was also a ∼63% increase in SSB consumption among US children of schooling age between the years 1989 to 2008^7^. Racial disparities in SSB consumption are also evident^8-10^. For example, the odds of drinking SSBs one or more times per day is significantly higher among non-Hispanic black respondents (AOR = 1.65) versus non-Hispanic white respondents^8^. This shift in increased SSB consumption and documented racial disparities are gravely concerning given the increasingly recognized role of excess dietary fructose in various, negative health outcomes^11-13^, including two recent landmark murine studies, which have begun to define mechanisms linking SSB/HFCS to colon carcinogenesis^14, 15^. Despite these studies, the role of dietary fructose in healthy human colon, and as such, the mechanisms that may drive fructose-mediated CRC risk, remain poorly understood.

An increasing body of observational studies has directly linked intake of fructose in obesity, diabetes and CRC risk^11, 16-18^. A small study in Iran recently found that increased fructose intake was associated with CRC after adjusting for multiple confounding factors including body mass index (BMI)^19^. This finding is in line with results from a larger study of two prospective US cohorts, which found that dietary fructose and SSB consumptions were both positively associated with incidence and mortality of proximal CRC^18^. Other research has shown that high SSB consumption was also associated with early-onset CRC in women^16^. Notably, African Americans (AAs) have higher incidence of CRC compared to any other racial/ethnic group^20^. AAs are also more likely to be diagnosed at a younger age and with poorer survival outcomes than EAs^20, 21^. Moreover, AAs are more likely to develop right-than left-sided colon cancer and more likely to develop right-sided colon cancer than EAs^22^. However, the mechanisms driving the observed racial disparities in CRC risk and outcomes and tumor sidedness are poorly understood.

DNA methylation (DNAm) represents one epigenetic mechanism through which gene expression patterns may be modulated to alter disease risk in response to environmental cues^23^. Aberrant DNAm is a hallmark of CRC^24, 25^, and differences in DNAm have also been found to be associated with fructose and HFCS in other tissues^26-28^. We have established a repository of right and left normal colon biopsies from a cohort of well-characterized average-risk AA and EA individuals^29^. In this cohort, we previously reported colon location and race specific DNAm differences, where we found that many of these differences occurred in regions known to be associated with CRC. Further analysis of this cohort has the potential to reveal novel insight into the biological mechanisms that drive CRC racial disparities and to help inform the development of tailored prevention strategies.

Here, we extend the results identified in our previous study to identify differences in DNAm associated with total dietary fructose in AA colon. We hypothesize that this will provide novel insight into the mode of action of fructose in the human colon, and that through the overlay of CRC-related publicly available data, we may contextualize these mechanisms in the framework of CRC risk.

## Materials and Methods

### Collection of dietary data for analysis

Clinical data were obtained from the medical records and pathology reports following colonoscopy. Behavioral and lifestyle information were obtained from a computer-assisted personal interview (CAPI) based on the NCI Colon Cancer Familial Registry Risk Factor Questionnaire^30^. The CAPI was administered by trained interviewer. Dietary fructose intake was estimated based on a self-administered food frequency questionnaire (FFQ).

### Sample collection and pre-processing

This study was approved by Case Western Reserve University/UHCMC and the University of Virginia Institutional Review Boards (IRB-HSR-21219). This study followed Strengthening the Reporting of Observational Studies in Epidemiology guidelines. The data generated exist as publicly available data on Gene Expression Omnibus: GSE151732. Pre-processing was carried out using the *R* package, *minfi* ^31^. Full details can be found in Devall et al., (2021)^29^. Preprocessing and analysis of three cancer cohorts was carried out as performed previously^32^.

### Preprocessing and statistical analysis of DNAm data

For our analysis of fructose, we accounted for the effects of technical variation on the array while retaining biological variation attributed to fructose by adjusting for both sentrix ID and position using the champ.runCombat() function of the R package, *ChAMP*^33, 34^. Beta values were logistically transformed prior to correction and were then returned for downstream regression analysis. For the analysis of fructose on DNAm, the following model was built:

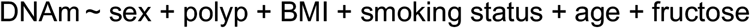

Sex = biological sex; polyp = polyp detected at any site along the colorectum; BMI = log-transformed BMI; smoking status = never/former/current; age = age at sample index; fructose = log-transformed measure of total dietary fructose intake.

Differentially methylated region (DMR) analysis was carried out using *DMRcate*^35^. The cpg.annotate() function was applied with the following arguments: what = “B”, fdr = 0.1, arraytype= “EPIC” and analysis.type = “differential”. Following annotation, the dmrcate() function was applied using the following arguments: C=4, lamda=1000, nCpG=3. Measures of false discovery rates (FDR) were calculated by applying the p.adjust() function to the resulting Stouffer P values for each region, as previously^32 36^. To identify DMRs of a similar minimum size, these same parameters were then applied to our repeated-measures analysis of three CRC datasets^25, 37, 38^, where we aimed to identified differences in DNAm between CRC tumor and matched, normal-adjacent tissue (NAT). Only one exception to our method was considered for this analysis: C=2 (default value for 450k data) when calling the dmrcate() function. Overlapping regions across datasets were defined using the in.region() and bedr.join.region() functions of the *bedr* package^39^.

### Statistical analysis of RNA-seq data

Bulk RNA-seq data was downloaded for The Cancer Genome Atlas Colon Adenocarcinoma (TCGA-COAD) dataset from *TCGAbiolinks*^40^. To identify differences in gene expression between matched (n=41 pairs) CRC tumors and normal adjacent tissues (NATs), we performed a repeated measures regression using *DESeq2*^41^.

## Results

### Dietary fructose consumption is associated with widespread DNAm differences in AA right colon

We used *DMRcate*^*35*^ to identify significant regions of DNAm that correlated with total dietary fructose consumption. Given the relatively limited sample size of EA individuals who provided data on dietary fructose consumption (n=34), we restricted our analysis AAs (n=79). In right colon, 4,263 DMRs survived FDR corrections. This included a hypomethylated DMR at chr1:9130738-9131854 (FDR=4.09E^-03^), a genomic region near the transcriptional start site of *SLC2A5* (**Figure 1A**). DNA hypomethylation of chr11:18415641-18415964, which corresponds to lactate dehydrogenase A (*LDHA*, FDR=0.036)) was also identified. Increased LDHA expression was previously identified in the intestinal epithelium of HFCS-treated mice^15^. To better determine the coordinated effects of dietary fructose on DNAm, we performed pathway enrichment analysis using *missMethyl*^*42*^. Here, we identified enrichments for terms previously associated with fructose, such as ether lipid metabolism (P=9.61E^-05^)^43^, arachidonic acid metabolism (P=1.56E^-04^)^44^ and linoleic acid metabolism (P=4.75E^-04^)^44^ (**Figure1B**). Other notable enrichments include the glycolytic process through fructose-6-phosphate (P=7.44E^-03^) and phosphofructokinase activity (P=9.53E^-03^), developmental process (P=6.20E^-04^), long-chain fatty acid metabolic process (P=5.45E^-03^) and cell population proliferation (P=9.10E^-03^).

**Figure 1:**
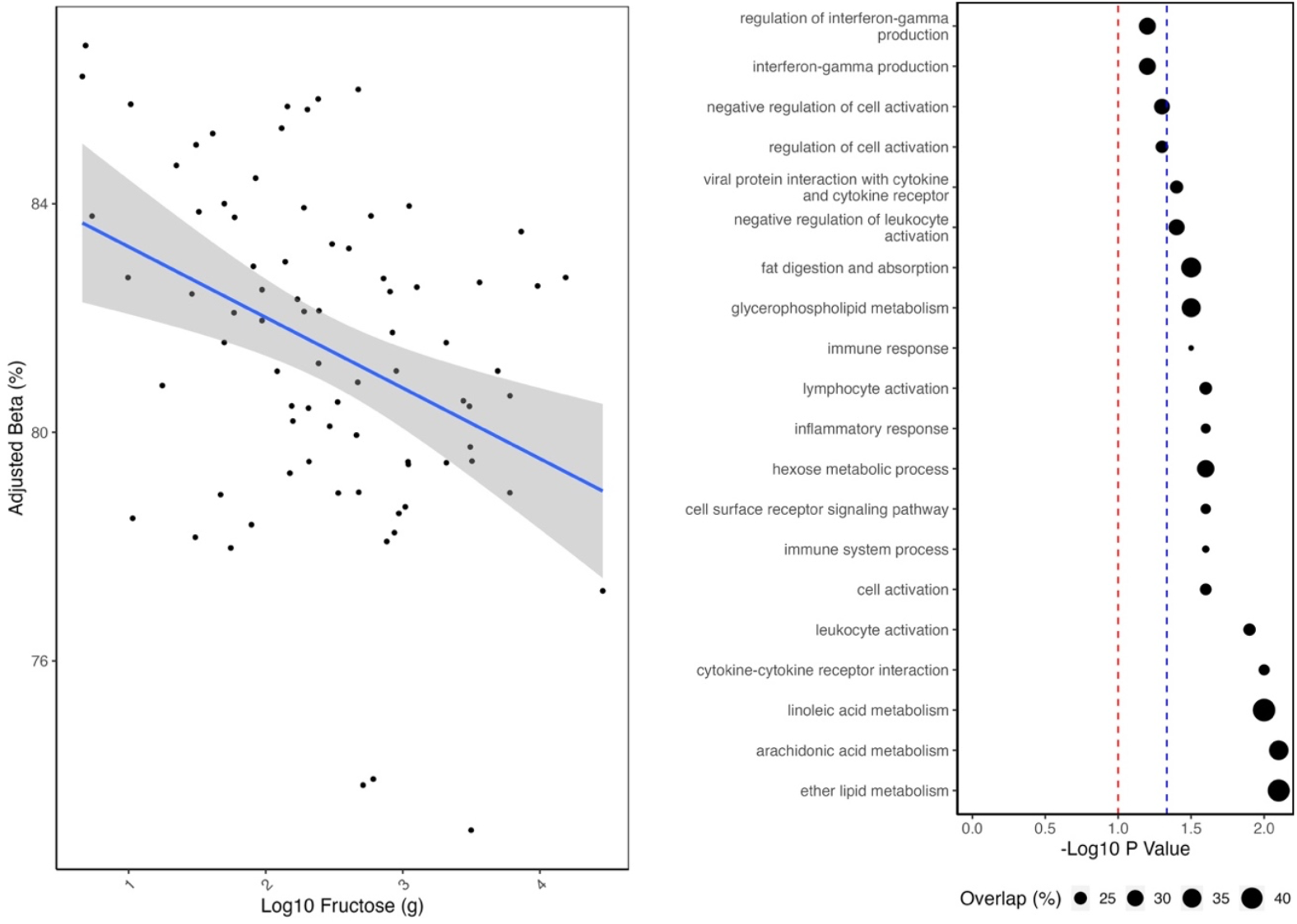
Summary of DMR analysis of dietary fructose in right AA colon. **(A)** *SLC2A5* was significantly hypomethylated in subject biopsies who consumed increasingly high amounts of fructose. **(B)** Summary of the top 20 most significantly enriched terms within our DMR analysis. “Overlap” corresponds to the number of DMRs identified within our analysis divided by the total number of genes within that pathway. Red and blue dashed lines correspond to nominal and FDR cut-offs, respectively.

### Dietary fructose consumption is associated with substantially fewer differences in differential methylation in AA left colon

In contrast to the widespread differential DNAm found to be associated with dietary fructose in the right colon, an analysis of matched, left colon biopsies revealed few associations. In total, 24 DMRs were associated with fructose in the left colon (FDR<0.05). Of the 85 nominal DMRs (P<0.05) associated with dietary fructose in the left colon, only 25 overlapped regions also found in matched, right colon. All 25 DMRs were consistent for direction of effect (**Table 1**).

**Table 1:** Summary of association findings through DMR analysis of fructose in right and left colon of AA. Findings were ordered from most to least significant based on the sum of their ranked significance. Bold font represents regions that were also identified in at least one CRC tumor analyses and displayed a concordant direction of effect.

| Gene | Right Colon |  |  |  | Left Colon |  |  |  |
| --- | --- | --- | --- | --- | --- | --- | --- | --- |
|  | Chromosomal Region | Beta<br>Diff (%) | P | FDR | Chromosomal Region | Beta<br>Diff (%) | P | FDR |
| **NBEA, MAB21L1** | **13:36050522-36051073** | **-0.046** | **1.16E<sup>-03</sup>** | **0.019** | **13:36050158-36051073** | **-0.040** | **1.22E<sup>-09</sup>** | **5.94E<sup>-07</sup>** |
| **ZFP30** | **19:38145827-38147309** | **0.005** | **7.93E<sup>-06</sup>** | **7.81E<sup>-04</sup>** | **19:38145827-38147309** | **0.006** | **1.67E<sup>-06</sup>** | **1.24E<sup>-04</sup>** |
| N/A | 11:128693801-128694679 | -0.023 | 1.55E<sup>-03</sup> | 0.023 | 11:128693677-128694679 | -0.029 | 4.48E<sup>-09</sup> | 1.56E<sup>-06</sup> |
| CRISP2 | 6:49681178-49681851 | -0.028 | 5.54E<sup>-03</sup> | 0.050 | 6:49681178-49681851 | -0.022 | 1.36E<sup>-07</sup> | 1.92E<sup>-05</sup> |
| FANCD2OS | 3:10149882-10150487 | -0.009 | 1.77E<sup>-03</sup> | 0.025 | 3:10149466-10150487 | -0.007 | 3.53E<sup>-06</sup> | 2.14E<sup>-04</sup> |
| MCPH1, ANGPT2 | 8:6419438-6419570 | -0.049 | 6.08E<sup>-03</sup> | 0.050 | 8:6418978-6420963 | -0.024 | 4.46E<sup>-07</sup> | 5.34E<sup>-05</sup> |
| CPT1B, CHKB-CPT1B, CHKB | 22:51016386-51017432 | -0.018 | 0.025 | 0.092 | 22:51016386-51017432 | -0.017 | 3.48E<sup>-10</sup> | 2.12E<sup>-07</sup> |
| TLE3 | 15:70371951-70372127 | -0.020 | 2.78E<sup>-03</sup> | 0.033 | 15:70371951-70373099 | -0.010 | 3.33E<sup>-04</sup> | 4.02E<sup>-03</sup> |
| N/A | 1:247537159-247537369 | -0.037 | 0.013 | 0.079 | 1:247537159-247537369 | -0.039 | 2.09E<sup>-05</sup> | 7.32E<sup>-04</sup> |
| ACSS3 | 12:81471311-81472177 | -0.025 | 5.20E<sup>-04</sup> | 0.015 | 12:81470331-81472177 | -0.015 | 7.26E<sup>-03</sup> | 0.022 |
| RP4-665J23.1 | 1:91300215-91300559 | -0.022 | 5.51E<sup>-04</sup> | 0.015 | 1:91300215-91300559 | -0.015 | 5.42E<sup>-03</sup> | 0.019 |
| N/A | 12:131865279-131865659 | -0.020 | 9.29E<sup>-03</sup> | 0.065 | 12:131864810-131865659 | -0.016 | 4.02E<sup>-04</sup> | 4.46E<sup>-03</sup> |
| **N/A** | **2:660692-660856** | **-0.020** | **0.023** | **0.092** | **2:660363-660856** | **-0.016** | **2.15E<sup>-04</sup>** | **3.20E<sup>-03</sup>** |
| HS3ST3B1 | 17:14213579-14214081 | -0.025 | 0.024 | 0.092 | 17:14213225-14214081 | -0.021 | 4.25E<sup>-05</sup> | 1.18E<sup>-03</sup> |
| DPP10 | 2:115419846-115420260 | -0.018 | 0.025 | 0.092 | 2:115419729-115420260 | -0.017 | 4.03E<sup>-05</sup> | 1.15E<sup>-03</sup> |
| AC093627.9 | 7:142966-143220 | -0.015 | 0.026 | 0.092 | 7:142411-143220 | -0.014 | 2.36E<sup>-04</sup> | 3.36E<sup>-03</sup> |
| ALS2CR11 | 2:202483740-202484320 | -0.015 | 0.049 | 0.115 | 2:202483740-202484583 | -0.013 | 4.37E<sup>-06</sup> | 2.51E<sup>-04</sup> |
| **N/A** | **14:73392078-73392648** | **-0.013** | **0.019** | **0.089** | **14:73392078-73392648** | **-0.006** | **4.19E<sup>-03</sup>** | **0.016** |
| **AL031663.1** | **20:44097309-44097731** | **-0.028** | **9.29E<sup>-03</sup>** | **0.065** | **20:44097309-44097563** | **-0.008** | **0.013** | **0.031** |
| N/A | 13:112212213-112212266 | 0.018 | 0.030 | 0.098 | 13:112212213-112212266 | 0.015 | 3.82E<sup>-04</sup> | 4.37E<sup>-03</sup> |
| RP11-23P13.6 | 15:42187036-42187219 | 0.022 | 0.019 | 0.089 | 15:42186830-42187219 | 0.014 | 0.013 | 0.031 |
| **N/A** | **14:52535758-52536436** | **0.019** | **0.034** | **0.100** | **14:52535758-52536721** | **0.014** | **5.63E<sup>-04</sup>** | **5.54E<sup>-03</sup>** |
| DTNA | 18:32173055-32173206 | -0.017 | 0.046 | 0.114 | 18:32173055-32173227 | -0.017 | 4.50E<sup>-04</sup> | 4.82E<sup>-03</sup> |
| WT1 | 11:32421514-32421845 | -0.020 | 0.032 | 0.099 | 11:32421514-32421845 | -0.015 | 2.53E<sup>-03</sup> | 0.013 |
| N/A | 10:131193104-131193272 | -0.019 | 0.050 | 0.115 | 10:131193104-131193272 | -0.014 | 2.19E<sup>-03</sup> | 0.012 |

### Contextualization of fructose-related findings in the context of CRC risk

We hypothesized that, if the fructose-related differences identified in our analysis contributed to CRC risk, then DMRs hypermethylated in response to increasing fructose would also be hypermethylated in CRC tumors. To address this hypothesis, we overlaid our findings with data obtained from a reanalysis of three CRC datasets^25, 37, 38^. Of the 4,263 right-side fructose-DMRs identified, 2,025 overlapped regions identified in at least one CRC tumor analysis and demonstrated the expected concordance for direction of effect. This included 1,373 DMRs that were present in all three CRC tumor analyses. In left colon, 28 (21: P<0.05, 7: FDR<0.05) fructose-related DMRs overlapped regions associated with CRC. Notable, nominal fructose risk DMRs of the left colon include chr13:28544471-28544820 (caudal type homeobox 2 (*CDX2*)), which was hypermethylated in both TCGA-COAD and ColoCare tumors and chr16:68676451-68676806 (cadherin 3 (*CDH3*)), which was hypermethylated in tumors of all three datasets. Of the 25 DMRs identified in both right and left colon, six were also identified in at least one CRC tumor analysis (**Table 1**, bold font).

### Defining DMR targets through analysis of TCGA-COAD

Given that some DMRs occur within intergenic regions or map near to multiple genes, we aimed to better define the likely targets of each fructose- and CRC-related DMR (fructose risk DMRs) by determining if their corresponding genes were found to be differentially expressed in CRC. Of the three cohorts considered in this analysis, only TCGA-COAD had matched RNA-seq data. A repeated measurement regression model was fitted for each gene to identify differences between matched tumor versus NAT across 41 pairs present within this dataset. Of the 1,581 fructose-related DMRs that were present in at least TCGA-COAD, 869 corresponded to differentially expressed genes (DEGs). Fisher’s test confirmed a highly significant enrichment for the differential expression of fructose risk DMR genes in TCGA-COAD (P=4.06E^-73^). In left colon, seven of 16 fructose risk DMRs with gene annotations corresponded to CRC DEGs (**Figure 2**).

**Figure 2:**
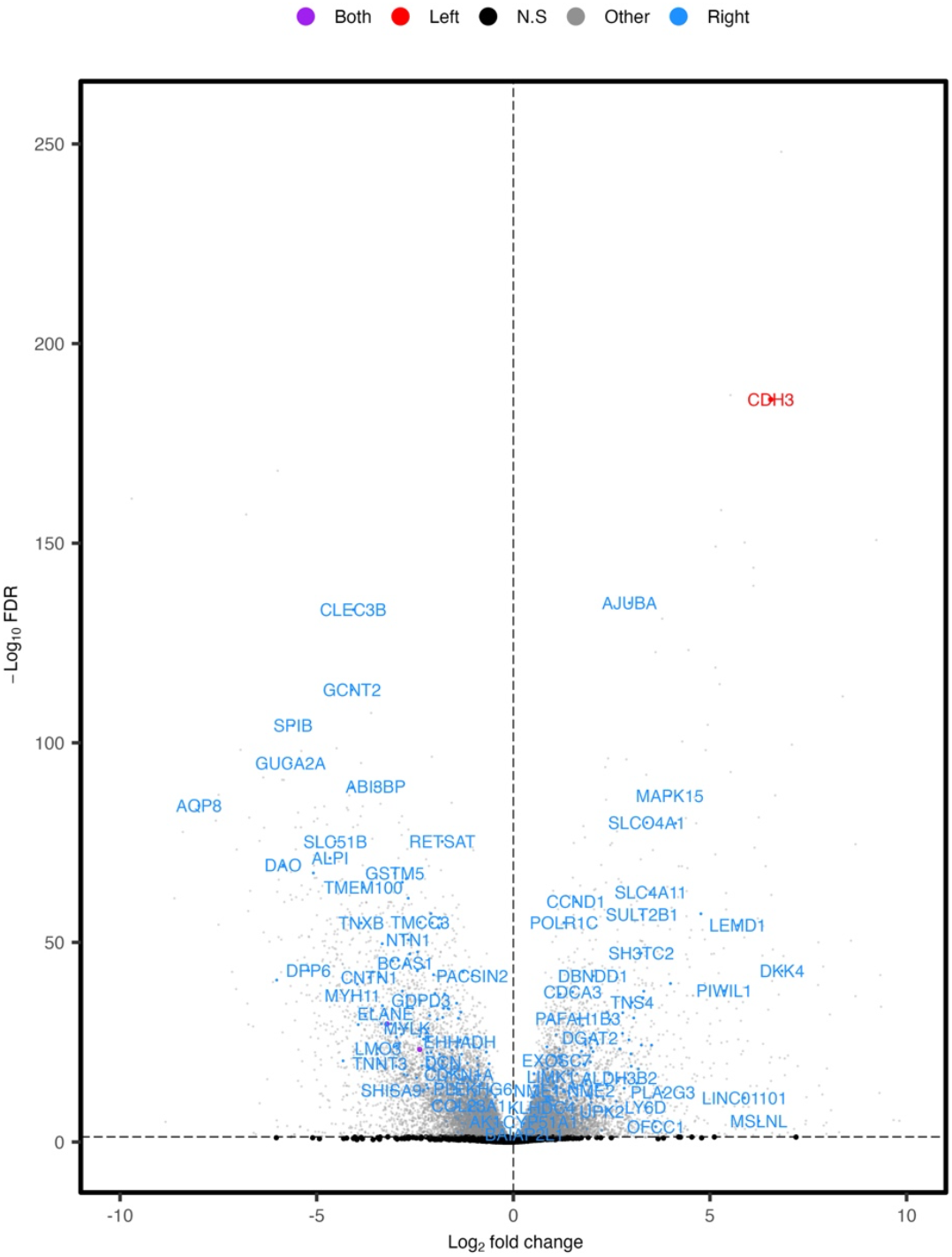
Volcano plot demonstrating the significance of genes corresponding to fructose-risk DMRs in TCGA-COAD expression data. Genes were color coded depending upon whether they were identified as fructose-risk DMRs in right (blue), left (red) or purple (both) analyses. Positive log_2_ fold changes correspond to DEGs overexpressed in CRC tumors versus matched NAT.

## Discussion

Our study represents the first, relatively large-scale interrogation of the association between DNAm and dietary fructose intake in normal colon tissues of AA patients free of CRC. It presents novel targets through which increased fructose consumption may drive CRC-related signaling. In recent years, there has been a growing interest in the use of dietary interventions to support cancer therapeutics^45^. Tumors frequently develop while retaining the metabolic pathways of their tissue of origin^15^. Cells that utilize the energy availability of their environment can display accelerated growth and outcompete their neighbors. Understanding the molecular drivers of these advantages in healthy colon tissue may therefore provide important mechanistic insight into the drivers of dietary risk genes. Such genes may not only provide biomarkers for risk, especially within individuals who consume high quantities of fructose, but may also provide novel targets for future therapeutics by focusing on genes driving preferential tumor-associated microenvironments.

Mouse studies have shown that intestinal absorption capacity is reached following moderate consumption of fructose (1g/kg), leading to unaltered fructose entering the colon^46^. Two landmark mouse model studies demonstrated a role for HFCS in colon carcinogenesis that was determined to be independent of its effect on body weight or the development of metabolic syndrome^14, 15^. They first show that HFCS enhances intestinal tumorigenesis through aberrant activation of glycolysis and promotion of fatty acid synthesis. Importantly, this study identified *KHK* as a key gene mediating this process. Later, Taylor et al., demonstrated that feeding mice a HFCS diet led to increased expression of hypoxia-inducible factor 1-alpha (HIF-1α), KHK and SLC2A5, which was accompanied by longer villi in both the proximal jejunum and the duodenum^15^. Further analysis in human CRC cell lines revealed that the metabolism of fructose by KHK led to the formation of fructose 1-phosphate, which promoted cell survival through the inhibition of another pyruvate kinase M2 (PKM2), thus contributing to cell survival under hypoxic conditions. These important studies were among the first to define a role for fructose/SSBs in CRC tumorigenesis. In the present study, our analysis of fructose risk DMRs did not identify any regions corresponding to *KHK*. However, we identified some overlap with the genes and pathways identified by both mouse studies. In this way, our human study provides partial support for previous findings in murine models, while also highlighting the imperative need for further mechanistic investigations into the role of CRC risk mediated by fructose in healthy, human colon epithelium. Several glycolysis and fatty acid terms were among the most significantly enriched in our analysis of fructose in right colon, and many of the DMRs that contributed to those terms, such as *CBFA2T3, HDAC4, HTRA2, PGAM2* (glycolysis) and *ACSF2, CYP2E1, PGIF, PLA2G4F, SLC27A5* and *TNXB* (fatty acid) overlapped regions identified in CRC tumors. Tumor cells frequently retain the metabolic pathways of their tissue of origin^15^ and the process of glycolysis provides an ample carbon source for fatty acid synthesis, which are required to house energy stores for the excessive energy demands of cancer proliferation^14^.

SSBs represent the leading source of added sugars in the US^47^ and added sugars account for over 25% of total calories for one in six AAs, in contrast to one in eleven EAs^48^, indicating that increased levels of dietary fructose consumption, particularly through SSB, may play a role. Importantly, AAs are 30% more likely to develop right-than left-sided colon cancer^21^, and are also more likely to develop right-sided colon cancer than EAs^49^. Differences in health care and dietary patterns driven by poorer socioeconomic status have been considered as contributory factors to observed disparities^22, 50^. While the role of SSB in racial disparities for CRC has yet to be fully explored, a previous multi-center study of 33,106 participants from the Nurses’ Health Study II found that SSB intake in adolescence was associated with increased adulthood adenoma incidence in proximal (right) colon and rectum. The same study also found that total fructose intake was only associated with increased risk for distal (left) and rectal adenomas^17^. However, a larger combined study of the Nurses’ Health Study and Health Professionals Follow-Up Study revealed that both SSB and total fructose were only associated with proximal/right-sided colon cancer^18^. Given that SSBs represent more of the daily caloric intake of AAs than EAs, they represent one potential driver of this racial disparities in CRC sidedness that warrants further investigation.

In line with findings showing a greater risk for dietary fructose in right than left colon cancer, our study identified a far greater number of differences in DNAm in right than matched left AA colon. Further, many of those DMRs identified exclusively in right colon (2,019 of 2,025) were also found in CRC tumors. Excess fructose enters the right colon following the saturation of SLC2A5 in the small intestine. Many of the DMRs identified in right colon were also differentially expressed in CRC tumors versus NAT and a subset (list subset) were also validated through a controlled dosing experiment of normal colon organoids. Our findings therefore imply a coordinated mechanism through which dietary fructose alters DNAm at regions relevant to CRC, leading to DEGs that modulate disease risk. The amount of fructose and its direct metabolites that are present in the left colon following further absorption to the hepatic portal system remains unclear. It is worth noting that while our study identified many more fructose-risk DMRs in AA right than left colon, a subset of fructose-risk DMRs exclusive to left colon were also found, including a hypermethylated DMR corresponding to the intestine-specific transcription factor, *CDX2*. A reduction in this tumor suppressor was also identified between CRC tumors and NATs in our analysis of TCGA-COAD. *CDX2* plays important roles in the regulation of epithelial cell differentiation and the reduction of its expression has been previously associated with poorer survival outcomes in left-sided colon cancer^51^. While the data presented within this study provides potential mechanisms that may drive the observed increase in right-sided colon cancer risk, larger epidemiological studies are needed to confirm this through finer site-stratified analyses. Given the notable differences in consumption of SSBs between AA and EA, and the relatively higher risk of AAs to develop right-sided colon cancer, these studies should also consider the effects of racial ancestry and ethnicity.

To relate our findings to CRC risk, we made use of publicly available CRC data and overlaid findings with those found to occur in CRC tumors. However, there are a number of limitations to this approach. First, CRC is a highly heterogenous disease and as such, it stands to reason that CRC risk pathways are also diverse. Second, total dietary fructose was highly correlated with other dietary sugars. Despite this, our results replicated multiple previously identified pathways and targets that are specific to fructose, including *SLC2A5*. However, validation through the coupling of high-throughput omics readouts and controlled dosing within the colon organoid system may further reveal mechanistic insight into the role of fructose in CRC. Nonetheless, our findings highlight several plausible targets through which dietary fructose may influence DNAm to modulate CRC risk and some of these targets were verified through fructose exposure to human AA colon organoids.

## Conclusion

There is increasing evidence for a role of dietary fructose and HFCS/SSB consumption in CRC risk. Here, we provide novel insight into colon location-specific mechanisms in AA subjects through which this risk may develop. That fructose consumption is associated with a far greater number of potential CRC risk DMRs in right than matched, left colon is intriguing. AAs are significantly more likely than EAs to consume greater quantities of fructose and to develop right-side colon cancer. Our findings highlight potential mechanisms through which increased dietary fructose consumption may differentially impact DNAm at regions associated with CRC to alter risk. Our findings provide some mechanistic support for reducing dietary fructose consumption.

## Data Availability

The data generated already exist as publicly available data on Gene Expression Omnibus: GSE151732, GSE101764 and GSE193535 and through the GDC Portal: TCGA-COAD.

https://portal.gdc.cancer.gov/repository?facetTab=cases&filters=%7B%22content%22%3A%5B%7B%22content%22%3A%7B%22field%22%3A%22cases.project.project_id%22%2C%22value%22%3A%5B%22TCGA-COAD%22%5D%7D%2C%22op%22%3A%22in%22%7D%2C%7B%22content%22%3A%7B%22field%22%3A%22files.data_category%22%2C%22value%22%3A%5B%22Transcriptome%20Profiling%22%5D%7D%2C%22op%22%3A%22in%22%7D%5D%2C%22op%22%3A%22and%22%7D&searchTableTab=cases

https://www.ncbi.nlm.nih.gov/geo/query/acc.cgi?acc=GSE151732

https://www.ncbi.nlm.nih.gov/geo/query/acc.cgi

https://www.ncbi.nlm.nih.gov/geo/query/acc.cgi

## Abbreviations

AA: African American
BMI: body mass index
CAPI: computer-assisted personal interview
CRC: colorectal cancer
DEG: differentially expressed gene
DNAm: DNA methylation
EA: European American
FDR: false-discovery rate
FFQ: food-frequency questionnaire
HFCS: high fructose corn syrup
NAT: normal-adjacent tissue
SSB: sugar sweetened beverage
TCGA-COAD: The Cancer Genome Atlas Colon Adenocarcinoma.

